## Supplementary Tables & Figures for "Microbial Sharing between Pediatric Patients and Therapy Animals during Hospital Animal-Assisted Intervention Programs"

### **Supplementary Materials**

**Supplemental Methods: Detailed DNA Extraction Protocol**

The Puritan swab samples were thawed, and 0.5 μL of Ready-Lyse Lysozyme (Epicentre Biotechnologies, Madison, WI) was added to each tube and incubated for 1 h with shaking at 600 rpm and 37°C. The swab was re- moved, placed into a spin basket, and centrifuged for 1 min at 9,400 × g to extract any remaining liquid. The sample was then added to a glass bead tube (0.5 mm; MO BIO, Carlsbad, CA) and vortexed for 10 min at maximum setting. The samples were then incubated in a heat block for 30 min at 65°C and 600 rpm, followed by ice for 5 min and a brief spin. A 150 μL of Protein Precipitation Buffer (Epicentre Biotechnologies, Madison, WI) was added, and the samples were vortexed briefly, then centrifuged at 22,000 × g for 10 min. The supernatant was removed and the protein pellet was discarded. The supernatant was mixed with 500 μL isopropanol and inverted to mix. The mixture was added to a spin column from the Genomic DNA Isolation Kit (Life Technologies, Grand Island, NY), and the remaining steps were followed according to manufacturer’s protocol. The samples were eluted with 50 μL Elution Buffer (Life Technologies, Grand Island, NY).

**Supplemental Figure 1: Study Design and Sampling Points**

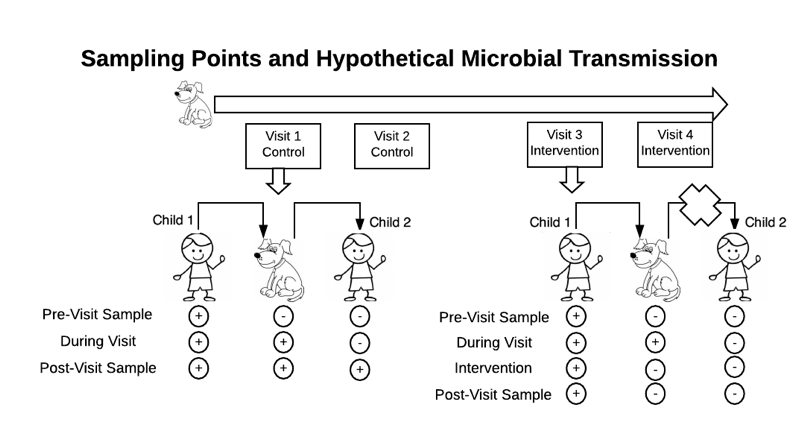

**Supplemental Table 1A: 16S rRNA gene Sequencing Library Results**

| **Read Counts** | | | | |
| --- | --- | --- | --- | --- |
| **Sample** | **Min** | **Mean** | **Median** | **Max** |
| Total | 1 | 10736 | 13342 | 21890 |
| Child N | 164 | 11639 | 14100 | 19430 |
| Dog All  N  MO  I  R | 711  711  4980  941  12745 | 12559  9315  14144  11389  15207 | 14448  8598  14989  13140  14967 | 21890  21890  16502  17373  18503 |
| Environment | 1 | 12051 | 13048 | 16109 |
| Field Blank | 520 | 4676 | 4307 | 10064 |
| Controls  Extraction  Sequence  Mock | 189  1  1 | 2357  26.2  4.5 | 2296  8  3.5 | 5207  117  10 |
| **Total DNA Concentration (ng/ul)** | | | | |
| Total | 0.04 | 8.517 | 3 | 41.39 |
| Child N | 0.05 | 5.434 | 2.25 | 31.67 |
| Dog All  N  MO  I  R | 0.16  0.16  0.6  0.18  1.49 | 11.001  3.92  18.8  3.19  16.9 | 7.05  0.975  19  2.14  13.9 | 41.39  18.5  41.1  9.46  41.39 |
| Environment | 0.07 | 19.77 | 19.98 | 38.82 |
| Field Blank | 0.2 | 0.7717 | 0.745 | 1.61 |
| Controls  Extraction  Sequence  Mock | 0.06  0.08  0.04 | 0.327  0.2267  0.105 | 0.31  0.16  0.08 | 0.67  0.7  0.22 |
| **qPCR 16S gene copies (/ul DNA)** | | | | |
| Total | 43 | 1.42e6 | 4570 | 6.26e7 |
| Child N | 111 | 16073 | 2470 | 2.58e5 |
| Dog All  N  MO  I  R | 208  208  1080  314  838 | 1.41e6  7630  1.72e6  10782  3.69e6 | 10850  1730  2.24e5  3940  95000 | 6.26e7  70100  2.52e7  77600  6.26e7 |
| Environment | 185e5 | 7.94e6 | 5.91e6 | 2.45e7 |
| Field Blank | 88 | 481 | 405 | 1040 |
| Controls  Extraction  Sequence  Mock | 56.6  61.9  42.8 | 460  344  233 | 273  293  181 | 1400  729  475 |

**Supplemental Table 1B: Microbial Community Sequencing Library Decontamination**

| Decontamination Stage | Number Contaminants (% removed) | Common Genera | Prevalence |
| --- | --- | --- | --- |
| Starting Taxa | **14183 total ASVs** |  |  |
| Sequencing Controls | 166 contaminants (1.17%) | *Corynebacterium*, *Sphingomonas*, *Streptococcus*, *Bacillus* | 832^nd^ most abundant |
| Extract Controls | 149 contaminants (1.06%) | *Corynebacterium*, *Streptococcus*, *Staphylococcus Sphingomonas* | 591^st^ most abundant |
| Field Blanks | 188 contaminants (1.36%) | *Corynebacterium, Streptococcus, Staphylococcus Sphingomonas* | 705^th^ most abundant |
| Final Taxa | **13680 total ASVs** (3.55%) |  |  |

Using ‘decontam’ package

**Supplemental Table 2: AAI Behavioral Observations and Patient-Therapy Dog Contact Scores**

| **Observed Interaction Behaviors** | **Patients Observed** |
| --- | --- |
| N * | 46 |
| Total time spent with dog, in minutes mean (range) | 14 (2-42) |
| Interact with Dog N (%) | 44 (95.7) |
| Sat on Floor N (%) | 28 (61.9) |
| Touch Head N (%) | 43 (93.5) |
| Touch Back N (%) | 27 (58.7) |
| Touch Belly N (%) | 13 (28.2) |
| Touch Paws N (%) | 11 (23.9) |
| Feed Dog N (%) | 24 (52.2) |
| Walk Dog N (%) | 10 (21.7) |
| Kiss Dog N (%) | 2 (4.3) |
| Hug Dog N (%) | 5 (10.9) |
| **Contact Score Level Calculation** | |
| Interact Score+ mean (range) | 7.89 (0-17) |
| Interact Score x Time ^  mean (range)  median (IQR) | 127 (0-403)  108 (56.25-197.75) |
| Contact Score Level  High (≥median) N (%)  Low (<median) N (%) | 25 (51%)  24 (49%) |

* observations not recorded for 3 patients

+ score based on total tally of individual behaviors, weighted based on closeness of contact: 1 point for interact with dog, walk dog, and sat on floor, 2 points touches, 3 points for feed, hug, or kiss dog.

^ score multiplied by total time with dog

**Supplemental Table 3: Relative Abundance by Host and Site by Phyla and Genus**

(Taxa > 3% total abundance)

| **Taxon (Phyla & Genus)** | **Patient Nasal** | **Dog Nasal** | **Dog Oral** | **Dog Perineal** | **Dog Inguinal** | **Environment** |
| --- | --- | --- | --- | --- | --- | --- |
| *Actinobacteria* | 0.0483 | 0.0330 | 0.0432 | 0.2035 | 0.0197 | 0.0522 |
| *Corynebacterium* | 0.1130 | 0.0330 | 0.0922 | 0.2035 | 0.0431 | 0.1968 |
| *Micrococcus* |  |  |  |  |  | 0.0313 |
| *Actinomyces* |  |  |  |  |  |  |
| *Bacteroidetes* | 0.0137 | 0.0213 | 0.0893 | 0.0704 | 0.0236 | 0.0248 |
| *Porphyromonas* |  | 0.0602 | 0.1604 | 0.1065 | 0.0673 |  |
| *Capnocytophaga* |  |  | 0.0883 |  |  |  |
| *Bacteroides* |  |  |  | 0.0734 |  |  |
| *Prevotella* |  |  |  | 0.0312 |  | 0.0365 |
| *Firmicutes* | 0.0667 | 0.1147 | 0.0212 | 0.0360 | 0.0341 | 0.0633 |
| *Staphylococcus* | 0.2714 | 0.2886 | 0.0333 | 0.0649 | 0.0453 | 0.0465 |
| *Streptococcus* | 0.1834 | 0.0411 |  | 0.0952 | 0.0432 | 0.1011 |
| *Faecalibacterium* |  |  |  |  |  | 0.0688 |
| *Blautia* |  |  |  | 0.0433 |  |  |
| *Alloiococcus* | 0.0382 |  |  |  |  |  |
| *Lactobacillus* |  |  |  |  |  | 0.0369 |
| *Dorea* |  |  |  | 0.0317 |  |  |
| *Fusobacteria* |  | 0.0120 | 0.0415 | 0.0370 | 0.0103 |  |
| *Fusobacterium* |  |  | 0.0415 | 0.0370 |  |  |
| *Proteobacteria* | 0.0368 | 0.0832 | 0.0559 | 0.0420 | 0.0191 | 0.0204 |
| *Moraxella* | 0.0572 | 0.1927 | 0.0396 | 0.0386 |  |  |
| *Conchiformibius* |  | 0.0462 | 0.1879 |  |  |  |
| *Lautropia* |  |  | 0.0741 |  |  |  |
| *Campylobacter* |  |  |  | 0.0648 |  |  |
| *Escherichia* |  |  |  | 0.0435 |  |  |
| *Pantoea* |  |  |  |  |  | 0.0427 |
| *Neisseria* |  |  | 0.0408 |  |  |  |
| *Sphingomonas* |  |  |  |  | 0.0349 |  |
| *Pseudomonas* |  |  |  |  | 0.0345 |  |
| *Spirochaetes* |  |  | 0.0274 |  |  |  |

**Supplemental Figure 2: Absolute Abundance of Key *Staphylococcus* Species**

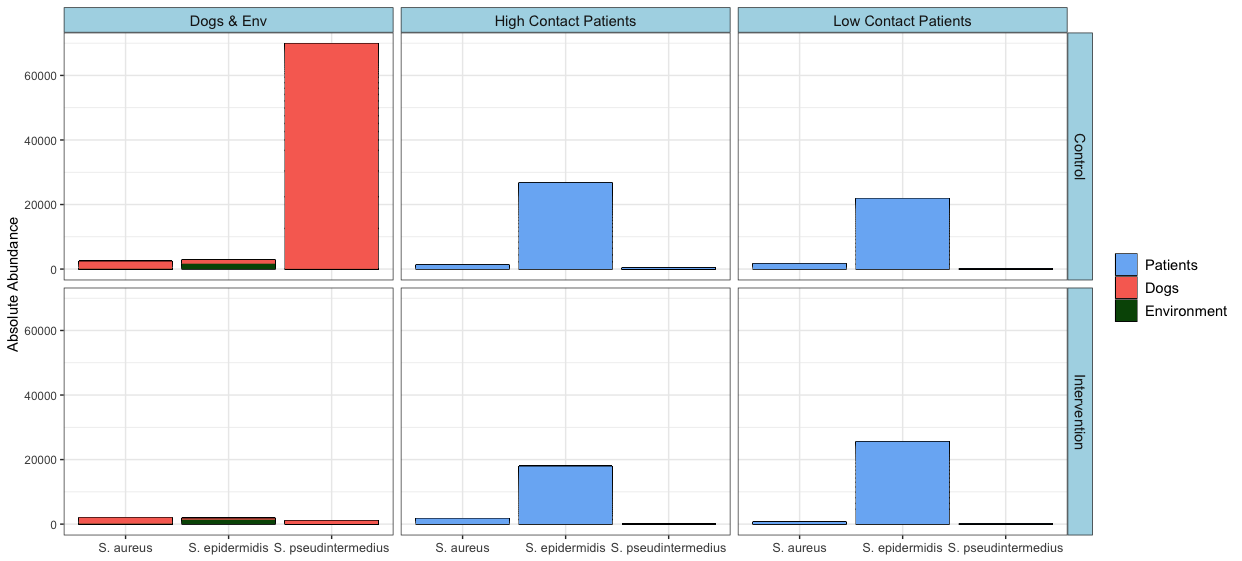

**Supplemental Figure 3: Alpha Diversity Curves by Host**

A. Total Taxa

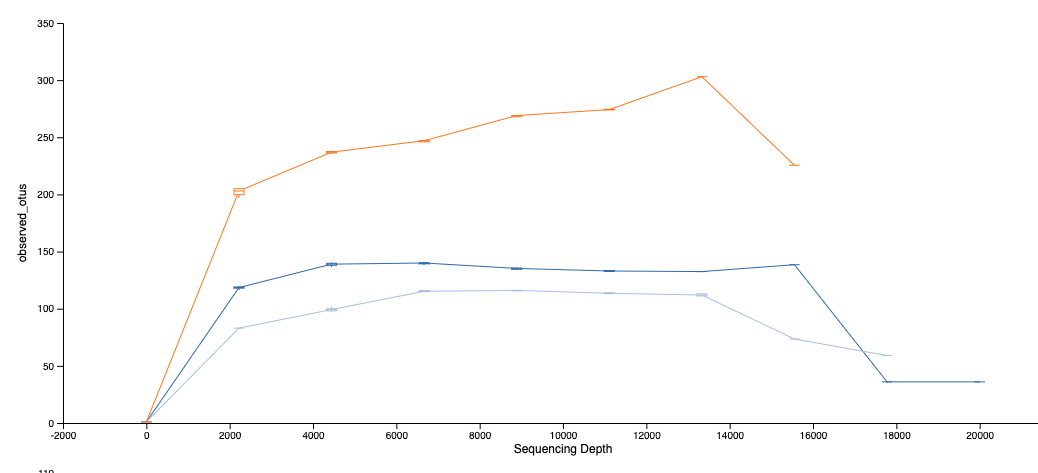

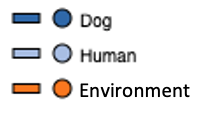

B. Shannon Diversity

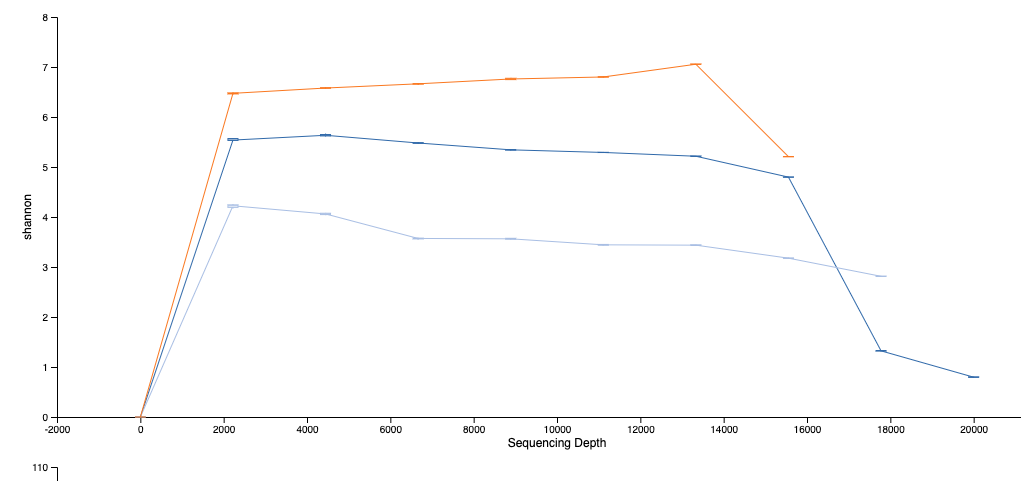

C. Faith’s Phylogenetic Diversity

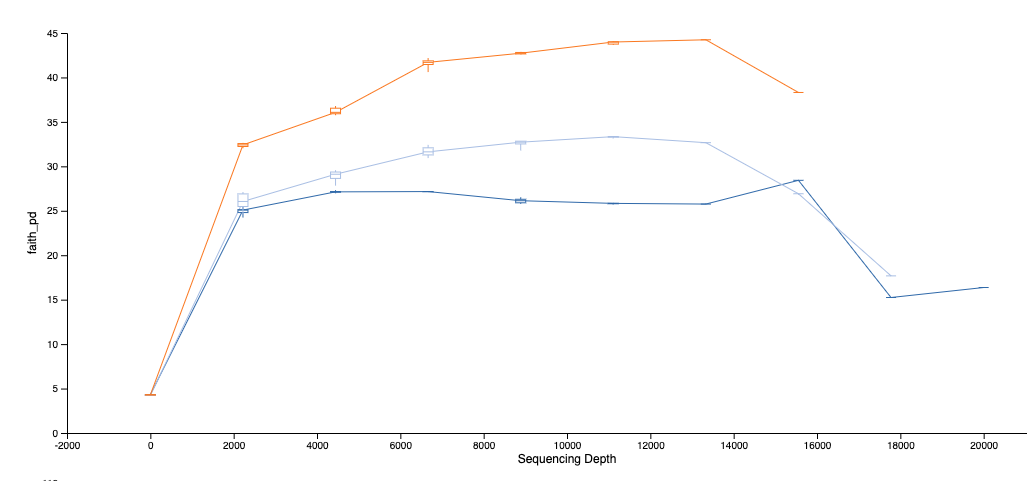

D. Number of Samples

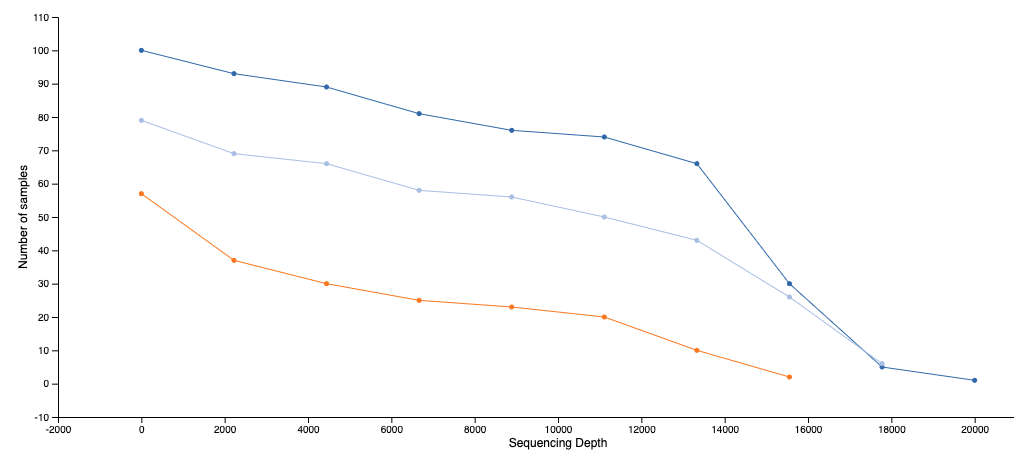

**Supplemental Figure 4: Beta Diversity Principle Coordinates Analysis Plots**

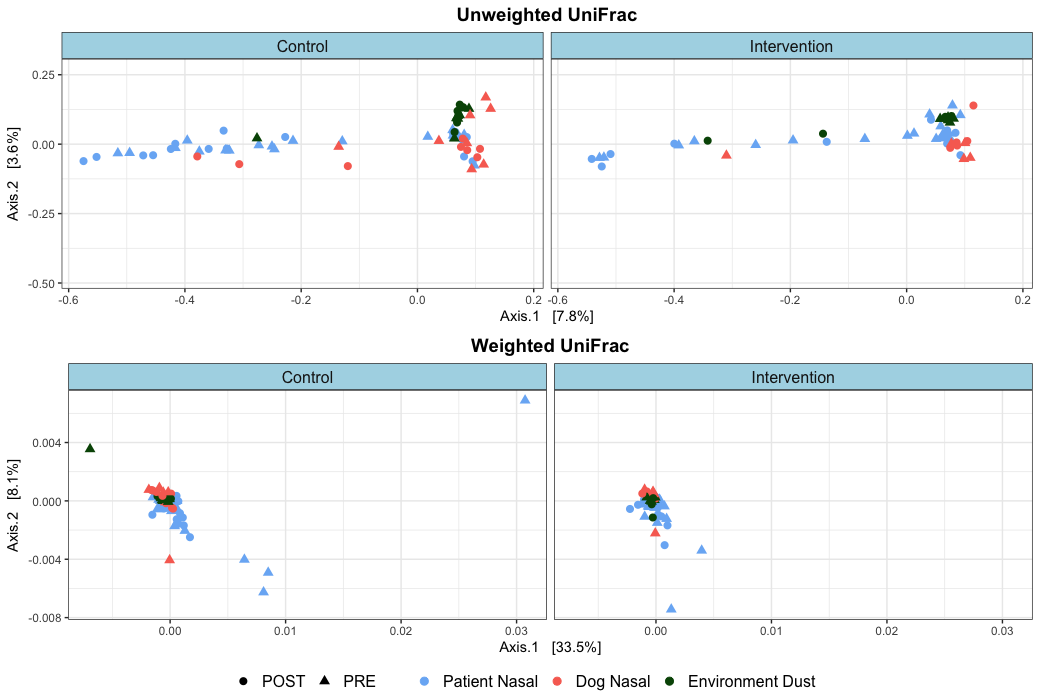

**Supplemental Figure 5: Graphical Example for Calculations of Beta Distance in Figure 4**

A. **Kid-Kid Distance Example**

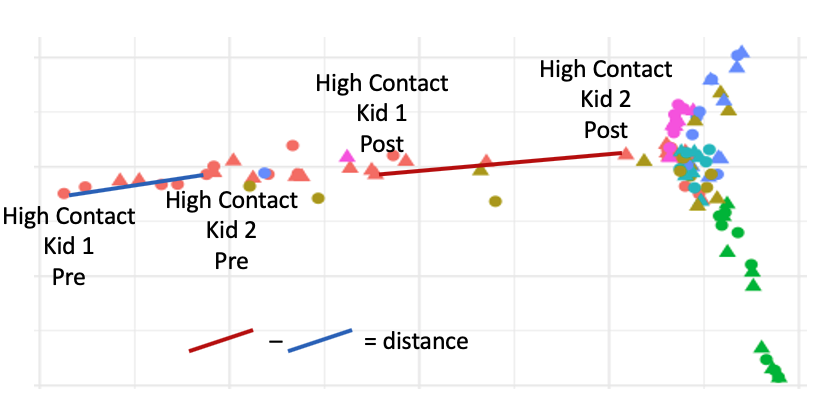

B. **Kid-Dog Distance Example**
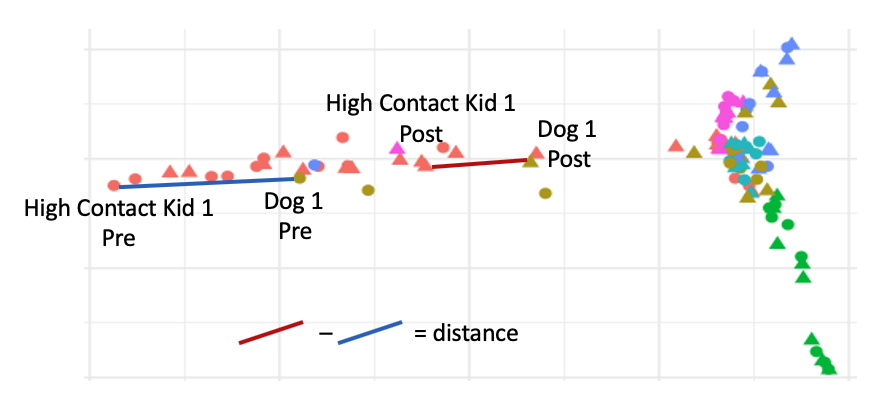

**Supplemental Figure 6: Visit Type and Contact Level in Microbial Composition Differences Between Patients and Between Patients and Therapy Dogs**

A. Unweighted UniFrac Distance

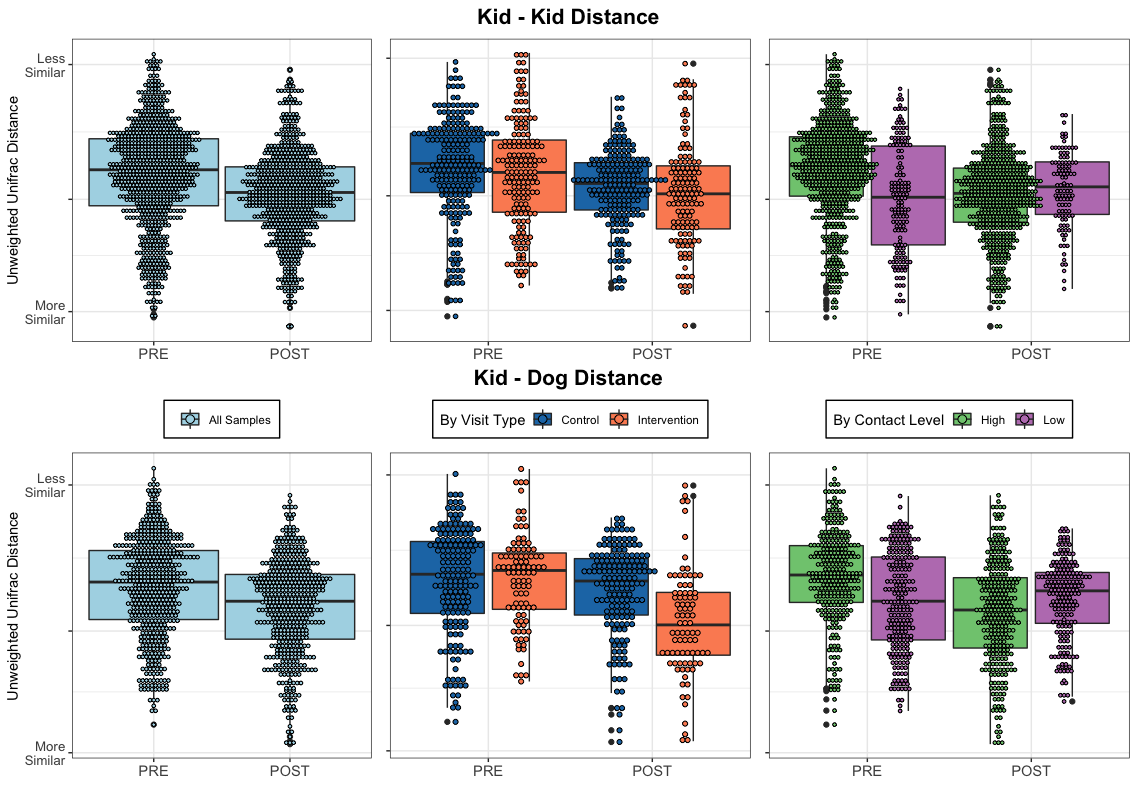

B. Weighted UniFrac Distance

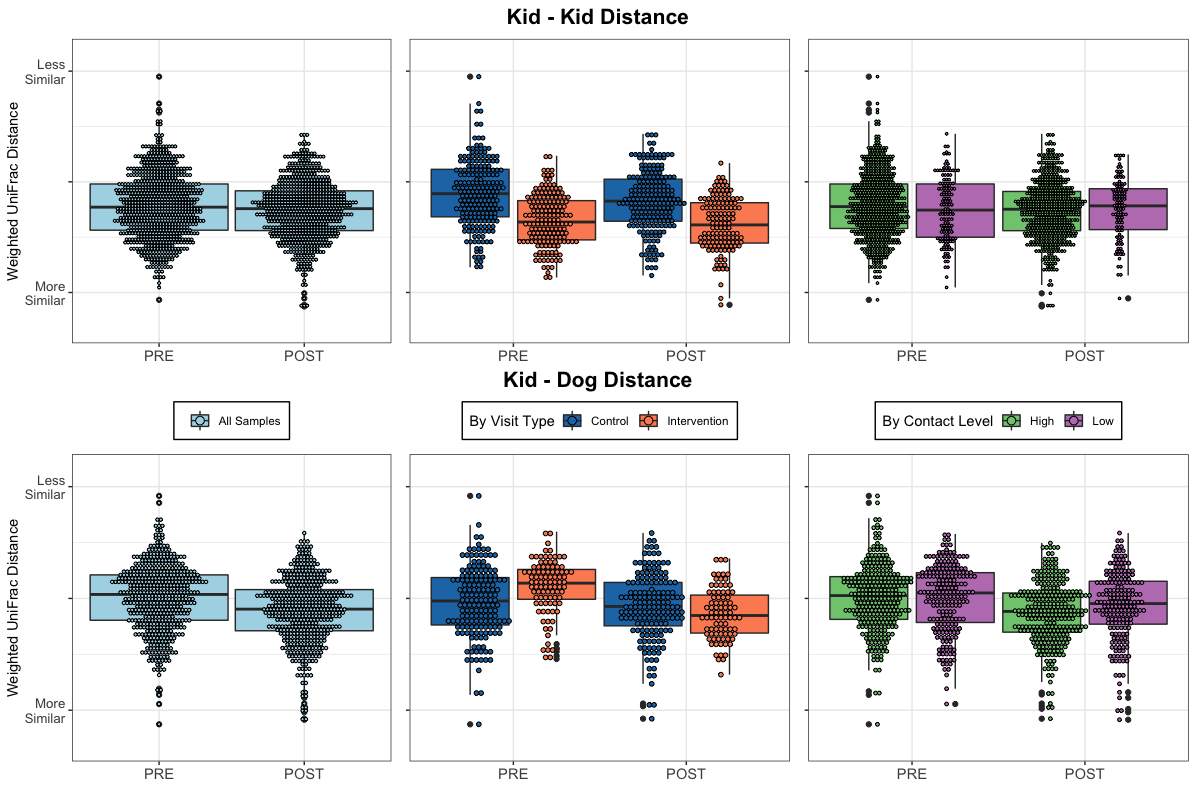

**Supplemental Table 4: Microbial Composition Differences between Patients and Between Patients and Therapy Dogs Before the Visit Compared to After Visit, Based on Visit Type and Contact Score**

Results for Beta Distance via PERMANOVA model

| **Group** | **Metric** | **Kid-Kid Distance** | | **Kid-Dog Distance** | |
| --- | --- | --- | --- | --- | --- |
|  |  | F-statistic | p | F-statistic | p |
| **Unadjusted Effect Pre versus Post** | | | | | |
| All Samples | Unweighted UniFrac | 47.068 | **0.0001** | 38.168 | **0.0001** |
|  | Weighted UniFrac | 19.33 | **0.0001** | 36.378 | **0.0001** |
| Control Visits | Unweighted UniFrac | 15.926 | **0.0002** | 2.294 | 0.128 |
|  | Weighted UniFrac | 17.78 | **0.0001** | 4.274 | 0.025 |
| Intervention Visits | Unweighted UniFrac | 8.737 | *0.003* | 35.188 | **0.0001** |
|  | Weighted UniFrac | 1.049 | 0.313 | 44.689 | **0.0001** |
| High Contact | Unweighted UniFrac | 89.538 | **0.0001** | 82.758 | **0.0001** |
|  | Weighted UniFrac | 16.017 | **0.0001** | 28.701 | **0.0001** |
| Low Contact | Unweighted UniFrac | 2.418 | 0.118 | 0.067 | 0.829 |
|  | Weighted UniFrac | 0.474 | 0.514 | 9.766 | *0.001* |
| **Multivariate Adjusted Effect** | | | | | |
| Pre versus Post visit | Unweighted UniFrac | 47.642 | **0.0001** | 38.626 | **0.0001** |
|  | Weighted UniFrac | 20.962 | **0.0001** | 36.422 | **0.0001** |
| Contact Level | Unweighted UniFrac | 10.317 | **0.0002** | 2.245 | 0.135 |
|  | Weighted UniFrac | 6.691 | **0.0003** | 0.829 | 0.328 |
| Visit Type | Unweighted UniFrac | 1.325 | 0.263 | 4.728 | 0.011 |
|  | Weighted UniFrac | 58.887 | **0.0001** | 2.138 | 0.094 |

PERMANOVA FDR-corrected **p<0.001**, *p 0.01-0.001*

Unadjusted Effect = difference in microbial composition distance between patients in pre-visit samples compared to microbial composition distance between patients in post-visit samples (kid-kid distance), or significant difference in microbial composition distance between patients and therapy dogs in pre-visit samples compared to microbial composition distance between patients and therapy dogs in post-visit samples (kid-dog distance) within each exposure group.

Example interpretation – In control visits, there is a significant difference in the microbial composition between patients before the visits compared to after the visit (p=0.0002 unweighted, 0.0001 weighted)

Adjusted Effect = independent effect of each exposure of the microbial composition distance between patients (kid-kid distance) or microbial composition distance between patients and therapy dogs (kid-dog distance).

Example interpretation – There is a significant difference in the microbial composition between patients with high contact compared to microbial composition between patients with low contact, independent of collection time (pre versus post) or visit type (control versus intervention) (p=0.0002 unweighted, 0.0003 weighted)
